# Changes in inflammatory and immune drivers in response to immunomodulatory therapies in COVID-19

**DOI:** 10.1101/2020.12.23.20248547

**Authors:** Stephen Y. Wang, C-Hong Chang, Matthew L. Meizlish, Parveen Bahel, Henry M. Rinder, Alfred I. Lee, Hyung J. Chun

## Abstract

As the global community strives to discover effective therapies for COVID-19, immunomodulatory strategies have emerged as a leading contender to combat the cytokine storm and improve clinical outcomes in patients with severe disease. Systemic corticosteroids and selective cytokine inhibitory agents have been utilized both as empiric therapies and in clinical trials. While multiple randomized, placebo controlled trials have now demonstrated that corticosteroids improve survival in patients with COVID-19,^1, 2^ IL-6 inhibition, which gained significant early interest based on observational studies, has not demonstrated reliable efficacy in randomized, placebo controlled trials.^3, 4^ To better understand the mechanistic basis of immunomodulatory therapies being implemented for treatment of COVID-19, we assessed longitudinal biochemical changes in response to such approaches in hospitalized patients with COVID-19. We demonstrate broad suppression of multiple immunomodulatory factors associated with adverse clinical outcomes in COVID-19 in patients who received corticosteroids, but no such response was seen in patients who either received tocilizumab or no immunomodulatory therapy. Our findings provide early insights into molecular signatures that correlate with immunomodulatory therapies in COVID-19 which may be useful in understanding clinical outcomes in future studies of larger patient cohorts.

---

Our analysis was approved by the institutional review board (IRB# 2000027792). Plasma samples were collected from patients admitted to Yale New Haven Hospital between 5/22/2020 and 5/27/2020 with COVID-19 infection (**Table 1**). Blood was collected on the day of hospital admission and on day 4 of hospitalization. Multiplex plasma profiling was conducted in patients who received no immunomodulatory therapy (n=4), tocilizumab (n=6), or corticosteroid (n=5) between day 1 and day 4 of their hospitalization. One patient in the corticosteroid group also received tocilizumab between the sample collection timepoints. Assays were performed by Eve Technologies (Calgary, Canada) through their Discovery Assay platforms. Fold change of biomarker levels were obtained by taking the log derivative of the ratio of the concentration of each factor on day 4 and day 1. Given the small sample size, our study was not designed nor powered to extrapolate clinical endpoints.

**Table 1.** Demographics and clinical outcomes of patients in the study.

| Patient | Immunomod. Treatment | Age** | Sex | Race | ICU (Y/N) | Intubation (Y/N) | Clinical Outcome (A=alive/D=dead) | Corticosteroid | Remdesivir (Y/N) |
| --- | --- | --- | --- | --- | --- | --- | --- | --- | --- |
| 1 | Neither | 45-50 | Male | Black | N | N | A | NA | N |
| 2 | Neither | 80-85 | Male | White | N | N | A | NA | N |
| 3 | Neither | 80-85 | Female | White | N | N | A | NA | N |
| 4 | Neither | 80-85 | Female | Black | Y | N | D | NA | N |
| 5 | Tocilizumab | 55-60 | Female | Black | N | N | A | NA | Y |
| 6 | Tocilizumab | 60-65 | Female | White | N | N | A | NA | N |
| 7 | Tocilizumab | 80-85 | Male | Black | Y | N | D | NA | Y |
| 8 | Tocilizumab | 55-60 | Male | White | Y | N | D | NA | N |
| 9 | Tocilizumab | 65-70 | Male | Black | Y | Y | A | NA | Y |
| 10 | Tocilizumab | 45-50 | Male | White | Y | Y | D | NA | Y |
| 11 | Steroid* | 60-65 | Male | White | N | N | A | Methylprednisolone | Y |
| 12 | Steroid | 50-55 | Male | Black | Y | Y | A | Methylprednisolone | Y |
| 13 | Steroid | 80-85 | Female | Black | Y | N | A | Methylprednisolone | N |
| 14 | Steroid | 40-45 | Female | Other | Y | N | A | Methylprednisolone | Y |
| 15 | Steroid | 70-75 | Male | Other | Y | Y | D | Methylprednisolone | Y |

\*Also received tocilizumab; \*\*Age range provided per MedRxiv policy

In general, the baseline levels of biomarkers in the three groups were not significantly different (**Figure 1**). In subjects receiving corticosteroid therapy, we found a broad suppression of multiple immunomodulatory factors that included a number of cytokines and chemokines that have been associated with adverse clinical outcomes in COVID-19 (IL-6, IL-8, IL-10, IP-10, G-CSF, MIG, MIP-1a, MCP-1, MCP-3, M-CSF and TNF-α) (**Figure 1)**.^5, 6^ Patients who received tocilizumab demonstrated the expected reciprocal increase in IL-6 levels seen with IL-6 receptor antagonism and had decreased levels of IL-1RA, IL-13, VEGF-A, and Angiopoietin 2 (Ang-2), but did not demonstrate reduction in the other COVID-19 related cytokines and chemokines which responded to steroid (**Figure 1**). Patients who did not receive either tocilizumab or corticosteroid showed an upward trend in IL-1b, M-CSF, and IL-10, and did not demonstrate any consistent decrease in COVID-19 related cytokines.

**Figure 1.**
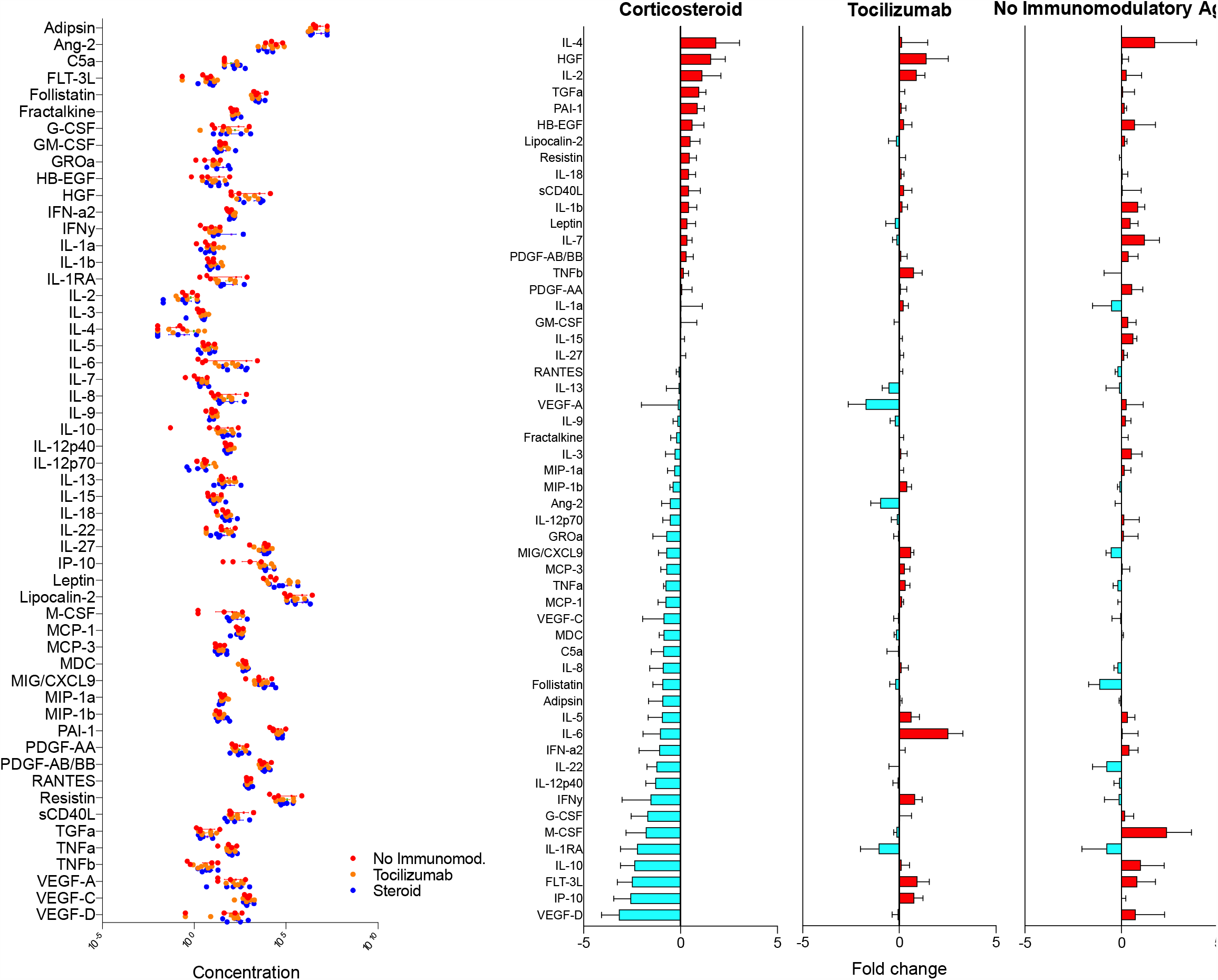
Comparison of circulating COVID-19 related biomarkers in response to immunomodulatory therapy. Left panel demonstrates baseline concentration of plasma biomarkers from hospitalized patients with COVID-19 are shown. All concentrations are in pg/mL except for C5a (complement 5a) which is ng/mL. Right panels depict changes in concentration of circulating biomarkers between days 1 and 4 of hospitalization grouped by corticosteroid (left), tocilizumab (middle), and no immunomodulatory therapy (right).

Our pilot study provides first broad, temporal profiling of patients treated with immunomodulatory therapies at the onset of their hospitalization for COVID-19. Therapeutic management of COVID-19 continues to vary across the world, with a paucity of robust mechanistic understanding to guide treatment choices. Our findings provide early insights into molecular signatures that correlate with the use of corticosteroid and tocilizumab which may be useful in understanding clinical outcomes in future studies of larger patient cohorts, and suggest that broader immunomodulatory strategy that targets multiple inflammatory pathways, rather than a single cytokine receptor, may afford maximal benefit in treatment of severe COVID-19. Furthermore, as other immunomodulatory agents are repurposed to combat COVID-19, research utilizing such immune biomarker-based approaches may provide a robust platform against which clinical outcomes can be correlated. These methodologies yield mechanistic insights that are urgently needed as the medical community aims to individualize COVID-19 therapies to maximize efficacy and minimize adverse effects.

## Data Availability

Primary data will be made available upon request.

